# Does neoadjuvant chemotherapy bias the transcriptomic profile of platinum-sensitive advanced high grade serous ovarian cancer patients towards a resistant phenotype? Findings from a gene expression meta-analysis

**DOI:** 10.1101/2025.09.23.25336416

**Authors:** Durga Prasan, Unnati Raut, Madhumathi HK, Gangotri Siddappa, Bonney LJ, Amritha Suresh, Sujan K Dhar

## Abstract

**Background:** Chemoresistance is the key determinant of long-term survival in advanced high-grade serous ovarian cancer (HGSOC). The molecular mechanisms underlying chemoresistance in clinical populations are not clearly known. The study aimed to analyze the molecular mechanisms promoting chemoresistance, as well as neoadjuvant chemotherapy (NACT) induced sensitive-to-resistant transformation in advanced-HGSOC patients.

**Methods:** RNA-sequencing data from Gene Expression Omnibus (GEO) database was systematically searched and extracted. Pre-chemotherapy (resistant vs sensitive) and chemosensitive (post-chemotherapy vs pre-chemotherapy) subgroup analyses were performed. For each subgroup, differential-gene-expression (DGE) meta-analysis and downstream analysis for pathway enrichment, protein-protein network and cancer stem cell expression were performed.

**Results:** Three bulk RNA-sequencing datasets (GSE162714, GSE173420, GSE227100) were included in the analysis. Prechemotherapy inherent-resistant samples showed upregulation of inflammation, epithelial-mesenchymal-transition processes, higher stromal proportion in tumors with upregulation of cancer-associated-fibroblasts (CAFs), and some of the cancer stem cell (CSC) markers over the sensitive phenotype. On the other hand, exposure to NACT in chemosensitive patients led to upregulation of inflammation, immune evasion, CSC-led proliferation and drug-efflux-pump overexpression, with increased immune and CAF cells in stroma mirroring an inherent-resistance phenotype, implying a sensitive to resistant transformation.

**Conclusion:** Inflammatory microenvironment, EMT and CSCs play a key role in promoting chemoresistance and post-NACT sensitive-resistant transformation.

## 1. INTRODUCTION

High grade serous ovarian carcinoma (HGSOC) remains one of the most challenging gynecological malignancies to treat. Despite being the third most common gynecological cancer by incidence, it has the highest proportional rate of mortality (1). Early peritoneal dissemination caused by shedding cancer stem cell (CSC)-rich spheroids from fallopian-tube epithelium into the peritoneal cavity distinguishes HGSOC from other epithelial ovarian cancers (2). The peritoneal microenvironment plays an important role in homing, survival, angiogenesis, and metastasis of CSC-rich spheroids, promoting disseminated peritoneal metastasis at initial clinical presentation (3,4). Surgical management of advanced HGSOC involves extensive peritoneal metastatectomy, called debulking or cytoreductive surgery. Residual disease after debulking surgery of peritoneal metastasis is an important determinant of survival (5). Taxane-platinum adjuvant chemotherapy remains the reference standard since early this millennium (6,7), forming an important adjunct to cytoreductive surgery (8,9).

Chemoresistance has been one of the strongest predictors of prognosis in HGSOC patients treated with chemotherapy (10). In fact, classification of platinum resistance is based on a survival parameter – the post-treatment platinum-free interval (PFI) of relapse (11–14). According to the 2010 Gynecologic Cancer Intergroup (GCIG) consensus statement, patients with PFI less than six months from the last platinum-based chemotherapy regimen are defined as chemoresistant, while others are considered candidates for platinum re-challenge (15). At presentation, nearly 70-80% of HGSOC patients are chemosensitive to platinum-taxane combination (16). However, with recurrent post-treatment relapses, a consistent rise in chemoresistance, leading eventually to the development of a chemorefractory state, is observed in nearly all the patients (17). Relapse free survival (RFS) period shortens progressively with each line of chemotherapy, implying a sensitive-to-resistant transformation induced by chemotherapy. This ultimately leads to extensive peritoneal and pleural dissemination of disease, bowel and respiratory morbidity, and eventual death (18). Five-year survival in advanced HGSOC has, thus, remained nearly static at 30% for the last three decades due to the development of chemoresistant relapses (19). Neoadjuvant chemotherapy-Interval Debulking surgery (NACT-IDS), which is an alternative to primary debulking surgery -adjuvant chemotherapy (PDS-ACT) protocol, improved surgical cytoreduction, but failed to improve disease-free or overall survival (20,21). An increase in the development of chemoresistant relapse could possibly contribute to the lack of expected survival benefit to NACT (22).

Understanding the mechanisms promoting chemoresistance is paramount to improving survival in advanced HGSOC. The emergence of antibody-drug conjugates (ADCs), where cytotoxicity is predominantly carried out by the chemotherapy drug payload, has further underscored the need to decipher chemoresistance mechanisms(23). Chemoresistance is either inherent, present at the initial untreated stage of disease, or acquired after chemotherapy. Nearly 15-25% of patients have inherent resistance, while the majority acquire it after chemotherapy exposure (acquired resistance) (24). Mechanisms underlying chemoresistance include cancer stem cells (CSCs) (25–27), epithelial-mesenchymal transition (EMT) (28,29), activation of drug efflux pumps(30), changes in drug absorption, modification of drug targets, epigenetic changes, altered metabolism, autophagy, and altered DNA repair mechanisms (31–33). Stroma and immune microenvironment have been increasingly shown to influence the development of chemoresistant milieu by promoting cell survival, angiogenesis, stemness, EMT and inflammation (34–36). Our understanding of molecular mechanisms in chemoresistance is predominantly derived from *in vitro* studies and animal models. Clinical studies to understand resistance mechanisms are confined to small studies, often in heterogenous patient populations. At the clinical level, we do not fully comprehend whether resistance is predominantly inherent, or if NACT contributes to the development of resistance, thereby nullifying the gains achievable by better cytoreduction. Patients treated with NACT-IDS protocol form excellent clinical models to study molecular mechanisms involved in chemoresistance. In this study, we performed extensive re-analysis of multiple publicly available gene expression datasets to understand critical pathways operating in chemoresistant patients. We also attempted to analyze pathways potentially involved in chemosensitive to resistant transformation induced by NACT.

## 2. METHODS

### 2.1 Patient Sample Datasets

Publicly available gene expression data was systematically searched following Preferred Reporting Items for Systematic Reviews and Meta-analysis (PRISMA) guidelines in Gene Expression Omnibus (GEO) database (**Fig 1A**). The search was focused on identifying high-throughput RNA sequencing datasets derived from pre- and post-NACT samples of Stage IIIc-IV HGSOC/ fallopian tube carcinoma/ primary peritoneal carcinoma patients treated with NACT-IDS protocol, published till February 29, 2024. Patients with other cancer histologies, early stage HGSOC treated with primary surgery and adjuvant chemotherapy, cell line or animal experiments were excluded from the search.

**Fig 1:**
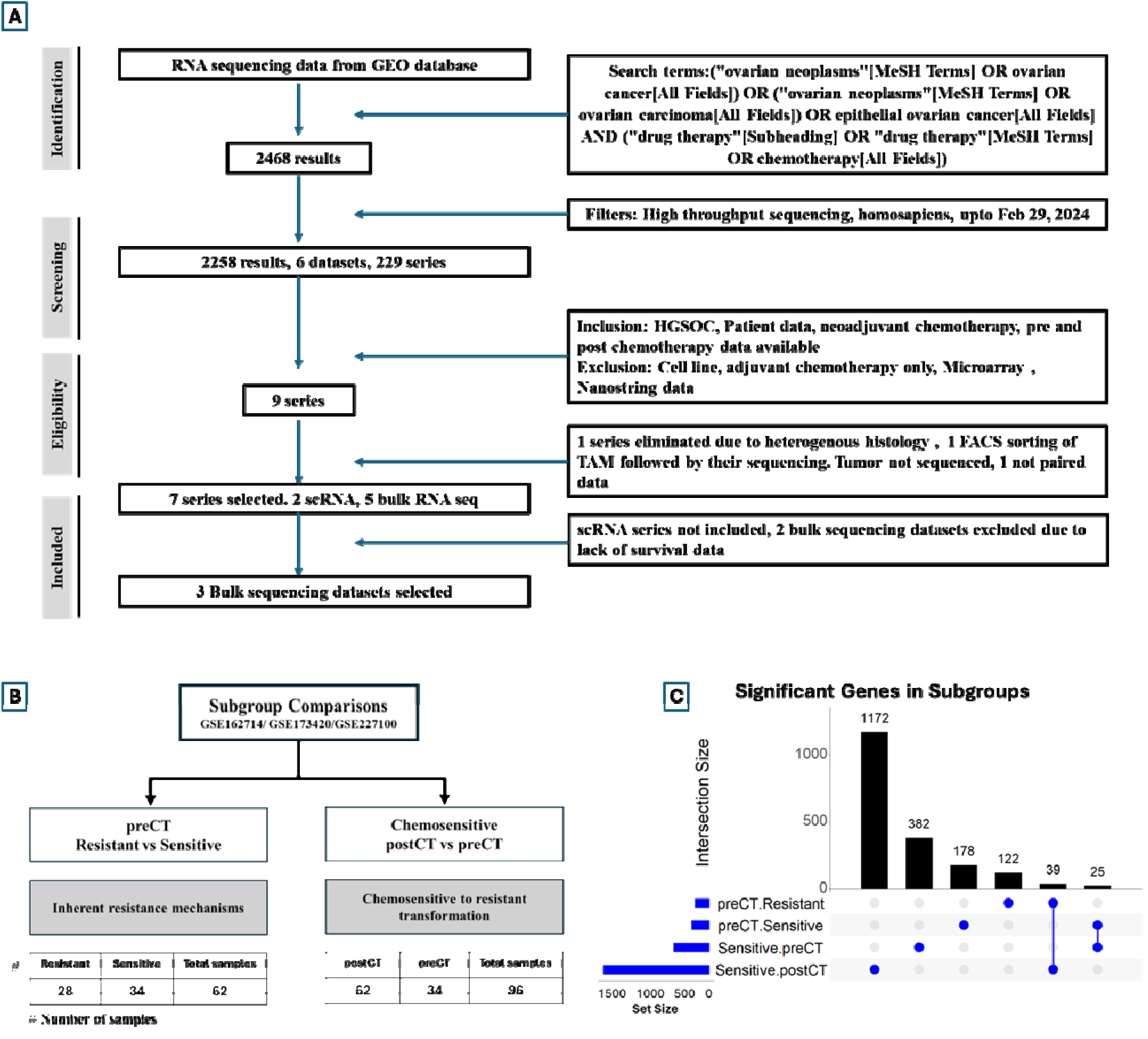
(A) PRISMA flow diagram for systematic search of GEO dataset. Out of 7 series which fulfilled the inclusion/exclusion criteria of stage IIIc-IV HGSOC patients treated with NACT-IDS treatment protocol and had pre and post chemotherapy tissue RNA-seq performed, the current analysis was performed on 3 bulk sequencing dataset which had metadata that enabled classification of patients into chemosensitive and chemoresistant groups based on relapse with platinum free interval of > or < 6 months respectively. Two bulk sequencing datasets were excluded due to lack of platinum free interval data (B) Summary of subgroup comparisons performed. The samples from 3 datasets included in meta-analysis was pooled and split based on preCT vs postCT, and chemoresistant vs chemosensitive. Our study looked at preCT (resistant vs sensitive) comparison to understand mechanisms distinguishing untreated inherent resistant from sensitive phenotypes, and Chemosensitive (postCT vs preCT) comparison to understand the mechanisms underlying chemosensitive to resistant transformation on administration of NACT. (C) Upset plot showing summary of number of significant genes upregulated on DGE in each sample group. The postCT group among Chemosensitive showed the highest number of genes upregulated (1172) implying widespread changes brought about by NACT administration overall. Thirty-nine genes were commonly upregulated among preCT resistant and chemosensitive postCT groups, signifying the markers common to inherent resistant samples and sensitive-resistant transformation.

Five bulk RNA-seq datasets (GSE71340, GSE162714, GSE227100, GSE173420, GSE227666) (37–42)were selected, out of which fastq reads for GSE173420 were acquired from the European Genome-Phenome Archive (EGA) with kind permission of University of Helsinki and Turku University Hospital Data Access Committee, while other datasets were downloaded from the NCBI Sequence Read Archive (SRA). All the datasets had reads generated from various Illumina sequencing platforms (**Table 1**). Reads were QC filtered using Trimmomatic, followed by alignment to the human genome GRCh38 reference using STAR and extraction of gene-level counts using the subreads featureCounts tool(43). Metadata from GSE162714 and GSE173420 contained survival and platinum-free interval (PFI) information. Based on PFI of less than six months, patients were labelled as chemoresistant, while the rest were labelled as chemosensitive. While GSE227100 lacked PFI data, since the metadata included information on sensitivity/resistance based on late (more than 6 months PFI) and early (less than 6 months PFI) recurrence, these three datasets were synthesized for meta-analysis. Metadata from GSE71340 and GSE227666 lacked chemoresistance, survival and platinum-free interval information, and hence were excluded from meta-analysis.

**Table 1:**
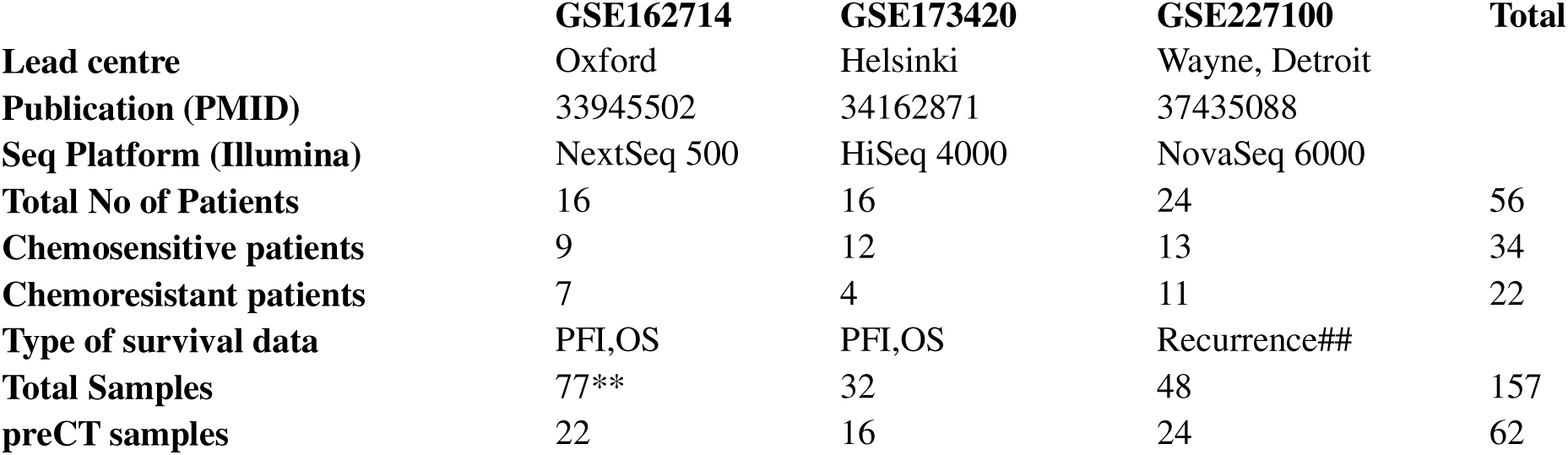
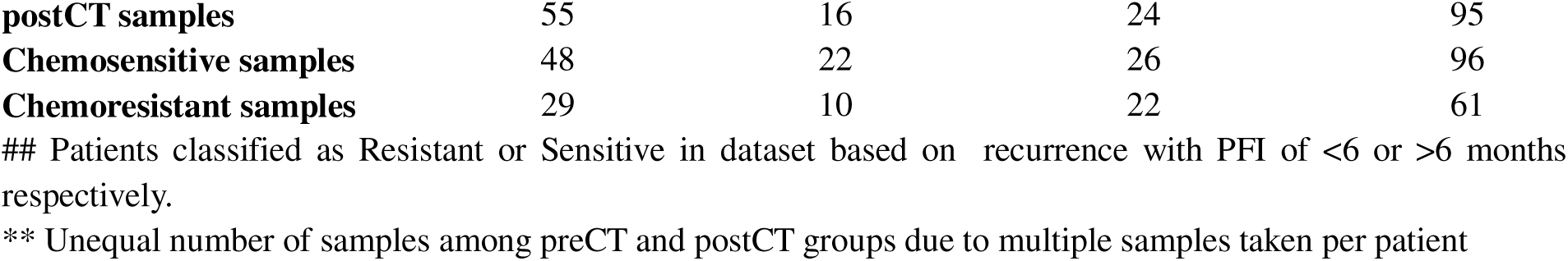
Details of datasets included in analysis.

Samples were divided into pre-chemotherapy (preCT) and post-chemotherapy (postCT) groups based on timing of sample collection, and were further categorized as Chemoresistant and Chemosensitive based on PFI criteria. This resulted in four subgroups for analysis, namely preCT (chemoresistant vs chemosensitive), postCT (chemoresistant vs chemosensitive), Chemosensitive (postCT vs preCT) and Chemoresistant (postCT vs preCT). The preCT subgroup provided insights into inherently resistant samples, while comparisons between preCT and postCT samples in the Chemosensitive subgroup probed the mechanisms involved in the transformation from sensitive-to-resistant induced by NACT. Our study was focused on these questions (**Fig 1B**).

### 2.2 Bioinformatic workflow

Within each subgroup, raw counts from multiple datasets were batch normalized using ComBat-seq function of sva R package (44). In order to ensure that malignant cell proportion in samples from each subgroup was adequate, Bayesian deconvolution of batch normalized counts was performed, and the distribution of malignant cells and stroma assessed in each subgroup comparison. Differential gene expression (DGE) analysis within subgroups was performed using DESeq2 (45). Results of individual DGE analysis were pooled in meta-analysis by Random Effects Model (REM) using MetaVolcanoR package (46). In order to broaden the analysis, significant DGEs were defined as absolute fold-change> 2.0 and Benjamini-Hochberg adjusted *p* value<0.05. Hub genes were identified using String network (47) with interaction score 0.4 and upto 5 interactors in the first shell, which were input into Cytoscape (48) and cytoHubba (49) plugin to obtain the top 10 upregulated genes by degree method using the shortest path.

Gene set enrichment analysis (GSEA) was performed for each analysis group using GSEA software (50). Significantly enriched pathways were defined at a false discovery rate (FDR)<0.25 and a nominal *p*-value of <0.10. Pathway overrepresentation analysis (ORA) was performed by filtering the markers from DGE results by p-value cutoff of <0.05, sub-setting the normalized count matrix for the selected markers, and clustering them by the bluster R package (51), which uses an ensemble of methods for cluster identification. The genes involved in the clusters were extracted and ORA done by the clusterProfiler R package(Yu et al., 2012) using Hallmark and Gene-Ontology biological processes (GO-BP). In order to identify fully interconnected core network, or cliques, of markers which could signify the most important biological processes in each subgroup, protein-protein interaction networks (PPIN) of markers from meta-analysis using a p value filter of <0.05 and absolute fold change > 2.0 were created separately using STRING (47) and PINOT (53) databases for each analysis group. The networks were combined to identify high-confidence interactions, where the combined score was greater than 0.5. Using the igraph R package, networks were built based on gene expression correlations (54). Immune cell deconvolution of batch-normalized RNA-seq counts was performed using immunedeconv R package (55), which uses ensemble methods for deconvolution. In addition, cell annotation was performed by Bayesian statistical analysis using ovarian cancer single cell signature in InstaPrism R package (56). Further, tissue-specific distribution in tumor microenvironment was identified using single cell dataset GSE165897, which compared pre- and post-chemotherapy gene expression at single-cell level from eleven patients with advanced HGSOC (57). Non-parametric test (Wilcoxon signed rank test) was used for comparing groups. Significance was defined by p-value <0.05. Details of software and versions are provided on the supplementary table.

### 2.3 Stem cell marker profile

In view of the important role played by cancer stem cells (CSCs) in the development of post - treatment relapse and chemoresistance, the current study laid specific focus on changes in CSC marker profile. The definition of CSC markers was done from a curated database (58) using a confidence score > 0.4, identifying 170 out of 8537 genes from the Biomarkers of Cancer Stem Cells database (BCSCdb). Changes in gene expression of these selected markers were noted using DGE meta-analysis results. In order to identify CSC markers with proliferative potential, significantly overexpressed CSC markers were queried in ovarian cancer cell-line datasets from BioGRID ORCS CRISPR screen database (59) and “hits” were identified.

## 3. RESULTS

### 3.1 Characteristics of datasets included

Selected bulk RNAseq datasets contained a relatively homogenous population of advanced HGSOC patients undergoing NACT-IDS based protocol with at least two chemotherapy drugs, taxane and platinum. All the patients received a minimum of three chemotherapy cycles before IDS. (**Table 1**).

The number of genes significantly differentially expressed in the subgroups, as well as the number of common markers is represented in **Fig 1C**. In chemosensitive samples, NACT exposure led to the highest number of DGEs, detailed below.

### 3.2 Inherent resistance versus sensitive

In preCT subgroup, both inherent-resistant and sensitive patients had an overlapping distribution of malignant cells and stroma-immune cells in the tissues sampled using Bayesian deconvolution (**Fig 2A & B**). The top ten hub genes identified in resistant samples by DGE meta-analysis and cytoHubba comprised cytokine ligands and receptors involved in immune reaction/ inflammation (CCL19, CCR6, CD69, CXCL5, MS4A1, GNG4, GZMK, PTGS2) and HOX family of genes implicated in EMT (HOXA3, HOXA2) (**Fig 2C**). Inflammatory response and epithelial-mesenchymal transition (EMT) were among the top five gene sets enriched on GSEA (**Fig 2D &E**). Resistant samples showed significant upregulation of CSC markers CXCL5 (**Fig 2F**), while the top 5 over-represented GO biological processes were related to chemotaxis, nitric acid metabolism, and inflammation (**Fig 2G**). Resistant samples were characterized by lower tumor purity/lower malignant epithelial cell proportion (Wilcoxon p=0.015), higher stromal (p=0.0096) and immune scores(p=0.051) on ESTIMATE cell type deconvolution (**Fig 2H**), and significantly higher fibroblast (p=0.012), plasma cells (p=0.011), B cells (p= 0.015) and mast cell (p0.035) populations (**Fig 2I&J**). Protein network cliques identified in resistant samples comprised of five markers involved (GZMK, CD69, CCL19, CCR6, MS4A1) in inflammatory reaction (**Fig2K**). In addition, DGE also showed significant upregulation of the HOX family of genes (HOXB9, HOXA10, HOXA4, HOXA2, HOXB6), which are associated with the mesenchymal TCGA subtype, possibly implying EMT as an important underlying process in resistance(**Supplementary File**).

**Fig 2:**
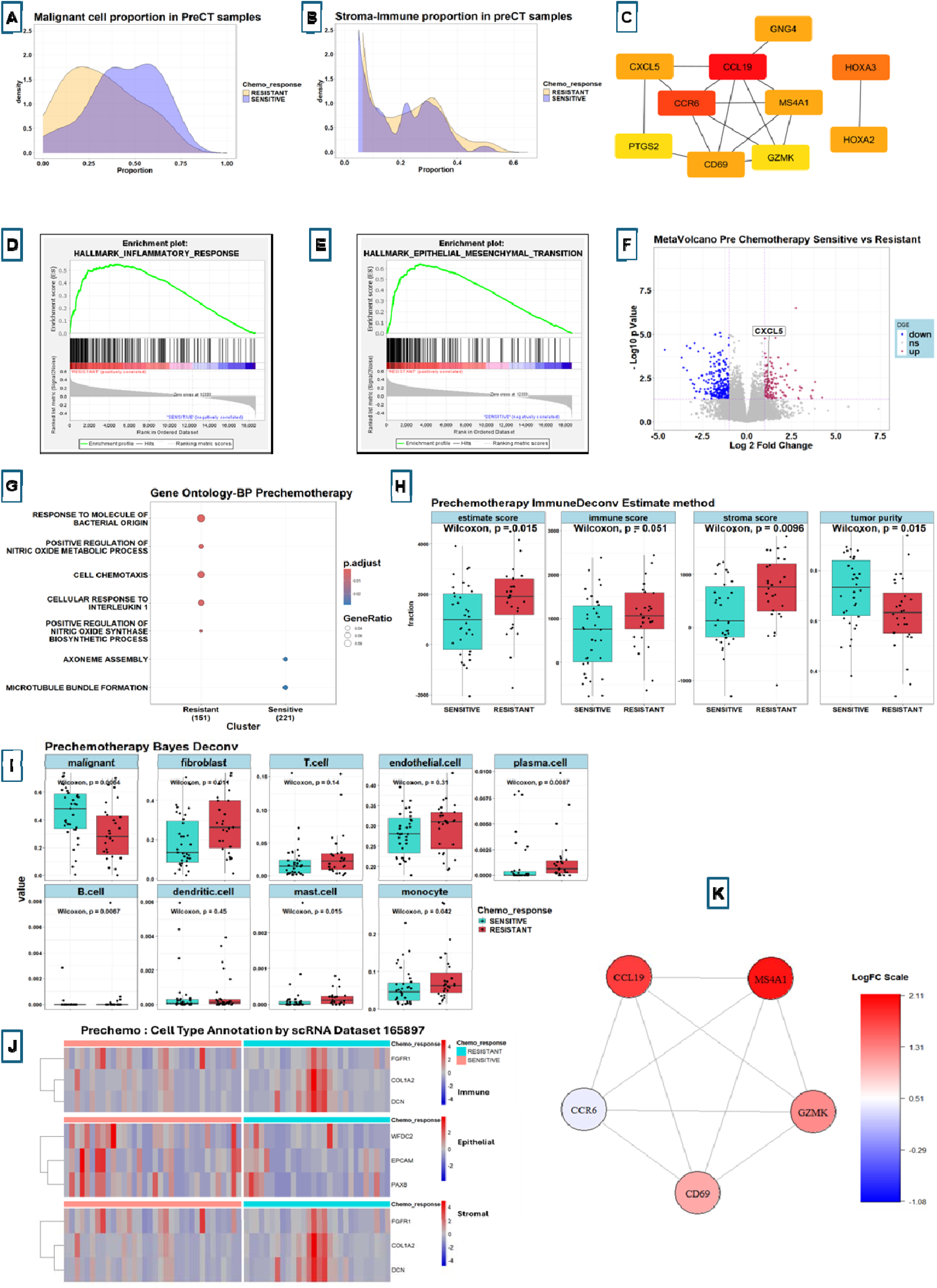
Summary of findings in Prechemotherapy (Resistant vs Sensitive) comparison. The distribution of malignant epithelial cells (A) and stroma-immune cells (B) among samples of both groups showing considerable overlap overall, suggesting similarity of sample quality in both groups. (C) Top 10 hub genes in inherent resistant samples showing genes involved in inflammatory response and EMT. Enrichment plots of Hallmark gene set GSEA showing significant upregulation of inflammatory response pathway (D) and Epithelial-mesenchymal transition (EMT) (E) in inherent resistant samples, both pvalue < 0.05. (F) Volcano plot of DGE showing significant upregulation of CSC markers (CXCL5) in inherent resistant samples. (G) Top 5 pathways upregulated in each group by ORA (Gene Ontology-biological processes) showing significant upregulation of inflammatory pathways and nitric oxide metabolism in inherent resistant samples. (H) ESTIMATE method of immune deconvolution showing significantly (p<0.05) lower tumor purity, and higher immune as well as stromal score in inherent resistant samples. (I) Bayesian deconvolution showing significantly lower epithelial and significantly higher fibroblast, plasma cells, B cells and mast cell population in inherent resistant samples. (J) Heatmap of cell annotation using GSE165497 single cell signature showing higher immune and stromal cells, and lower epithelial cell distribution in inherent resistant samples. (K) Fully connected network (clique) of genes generated using graph theory in inherent resistant samples shows 5 gene clique involved in inflammatory response.

### 3.3 Chemosensitive samples pre and post NACT

Among Chemosensitive patients, the distribution of malignant and stromal-immune cells in both precT and postCT samples was similar (**Fig 3A &B**), likely due to the quality of tumor sampling post-NACT in the included studies. Top ten hub genes postCT were inflammatory markers (CD4, CD8A, IL6, IFNG, IL2, PTPRC, CCR7, CD19, CD2, IL7R) (**Fig 3C**). Inflammatory response and TNFα signaling via NFκB were among the top five gene sets enriched on GSEA, though none reached significance (p>0.05) (**Fig 3D &E**). Chemotherapy exposure led to widespread changes in CSC marker expression, both upregulation and downregulation. Among the upregulated markers were known ovarian CSC markers (ALDH1A1), embryonic and CSC marker (KLF4, GLI1), ABC transporters involved in drug efflux pump mediated chemoresistance (ABCB1, ABCG2, ABCB5), proliferative markers (KIT, NGFR, RSPO2, CLU) and markers associated with inflammation (CTSG, IKZF3, SIRPG, IL33) (**Fig 3F**). Among them, ALDH1A1 and KIT had proliferative potential in BioGRID-ORCS CRISPR database. The top 5 pathways over-represented in the postCT cohort were related to inflammatory processes, while cell division-related processes were downregulated (**Fig 3G**). Deconvolution by the ESTIMATE method postCT showed a significantly lower tumor purity (p=0.005), and higher immune (p=0.0011) and stromal (p=0.035) score, findings which closely mimic inherent resistant phenotype (**Fig 3H**). All the immune cells (T cells (p=0.01), endothelial cells (p=0.0044), plasma cells(p=4e-04), B cells (p= 5.7e-0.5), dendritic cells (p=0.00023), mast cells 9p=0.0087), monocytes (p=0.003)) showed significant enrichment postCT (**Fig 3I**). Similarly, multiple cliques were identified in postCT samples, the largest being 20 marker clique comprised of markers involved in immune evasion and inflammation, implying the central role played by immune evasion possibly in sensitive to resistant transformation (**Fig 3K**).

**Fig 3:**
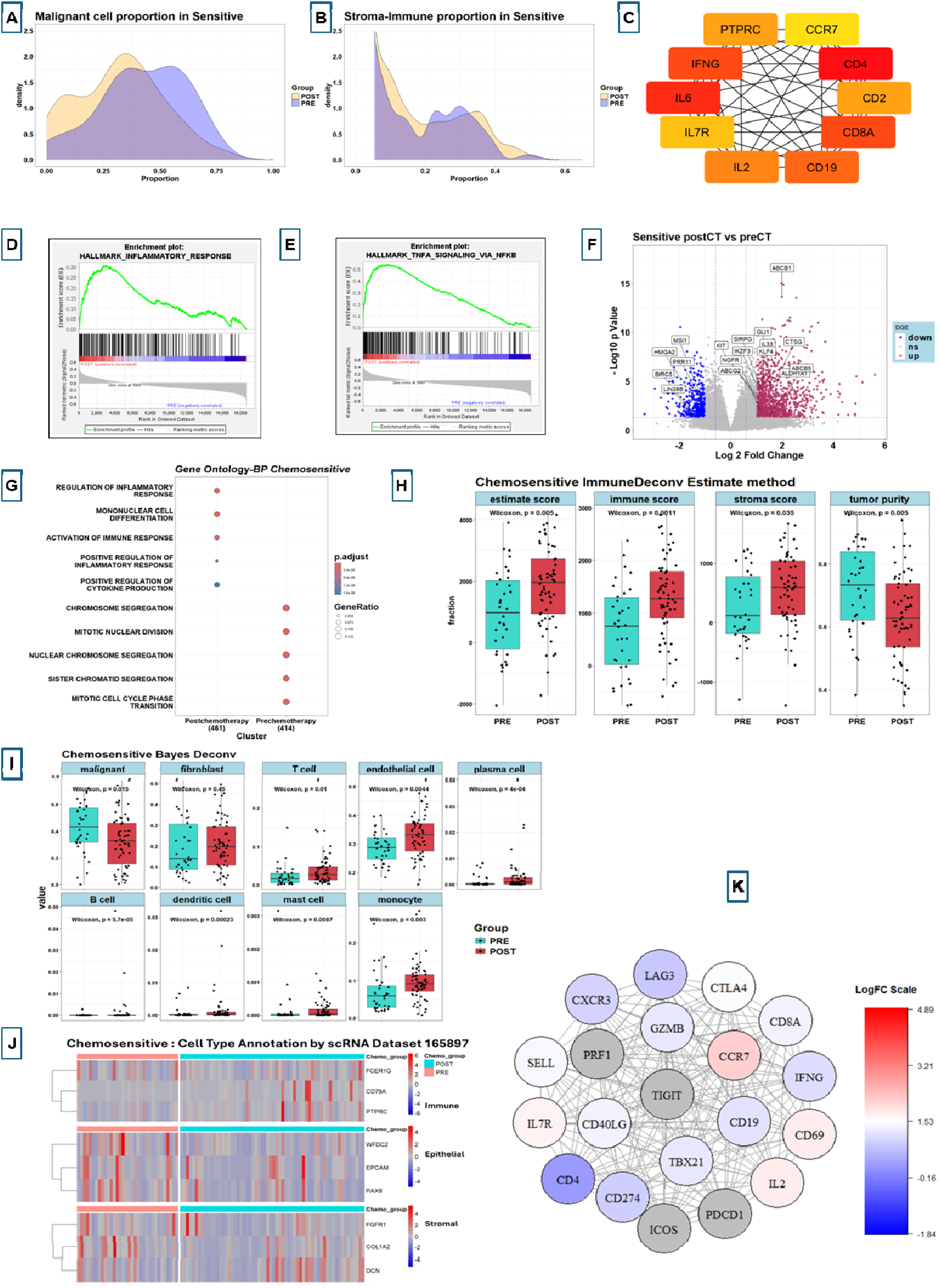
Summary of findings in Chemosensitive (postCT vs preCT) comparison. The distribution of malignant epithelial cells (A) and stroma-immune cells (B) among samples of both groups showing considerable overlap overall, suggesting similarity of sample quality in both groups. (C) Top 10 hub genes in postCT samples showing genes involved in inflammatory response. Enrichment plots of Hallmark gene set GSEA showing upregulation of inflammatory response pathway (D) and TNFA signaling via NFKB (E), both pvalue > 0.05, ns. (F) Volcano plot of DGE showing significant upregulation of CSC markers (ALDH1A1, KLF4, GLI1, ABCB1, ABCG2, ABCB5, KIT, NGFR, CTSG, IKZF3, SIRPG, IL33). (G) Top 5 pathways upregulated in each group by ORA (Gene Ontology-biological processes) showing significant upregulation of inflammatory pathways in postCT samples. (H) ESTIMATE method of immune deconvolution showing significantly (p<0.05) lower tumor purity, and higher immune as well as stromal score in postCT samples. (I) Bayesian deconvolution showing significantly lower epithelial and significantly higher immune cell (T cell, B cell, Dendritic cell, Mast cell, Endothelial cell, monocyte) cell population in postCT samples. (J) Heatmap of cell annotation using GSE165497 single cell signature showing higher immune and lower epithelial cell distribution in postCT samples. (K) Fully connected network (clique) of genes generated using graph theory in Chemosensitive postCT samples shows 20 gene clique involved in inflammatory response and immune evasion suggesting that immune evasion could be at the centre of sensitive to resistant transformation.

Predictably, NACT exposure in Chemosensitive samples led to widespread significant upregulation of ABC transporter genes (ABCA9, ABCA10, ABCA8, ABCB1, ABCA6, ABCD2, ABCB5, ABCC9, ABCG2, ABCC6), implicated in development of chemoresistance (**Supplementary File**). Interestingly, postCT samples also showed upregulation (fold change > 1.5) of immune evasion markers (CD274, TIGIT, CTLA4, LAG3) hinting at their possible contribution in sensitive—resistant transformation(**Supplementary File**).

## 4. DISCUSSION

Resistance to taxane-platinum chemotherapy has remained one of the critical determinants of survival in advanced HGSOC. The majority of patients with HGSOC are sensitive to this combination early in the course of the disease, but develop resistance eventually due to recurrent relapses and repeated exposure to chemotherapy. Our study is perhaps the first gene expression meta-analysis in advanced HGSOC patients undergoing NACT, which looks at a broad overview of resistance mechanisms. We selected a relatively homogenous population of patients diagnosed with HGSOC stages IIIc-IV and treated with taxane-platinum-based NACT-IDS protocol to avoid histology, stage or treatment-based confounding factors. TCGA datasets were excluded from our analysis since they lacked samples from NACT patients with pre and post-chemotherapy data.

### 4.1 Chemoresistance model in HGSOC

Based on our observations, we hypothesize two models for the onset of chemoresistance. In chemo-naïve patients presenting with an **inherent resistant** phenotype, cancer stem cells under the influence of CXCL5 produce a tumor microenvironment that is rich in stroma and immune cells, and lower epithelial cell purity. The epithelial cells undergo epithelial-mesenchymal transition promoted by HOX genes, as well as CXCL5 secreted by cancer-associated fibroblasts (CAFs) in the microenvironment. The HOX family is a master regulator of transcription and has recently been implicated in cisplatin resistance by negative regulation of cell differentiation (60). CXC and CC-cytokine ligands also induce an inflammatory microenvironment which, in combination with EMT, leads to an inherent resistant phenotype (**Fig 4A**). CXCL5 plays an important role in chemotaxis and suppression of CD8+ T cell-mediated immune responses to cancer in the microenvironment of lung cancer, thereby promoting tumor survival, angiogenesis, and resistance to treatment (61). CXCL5 plays diverse roles when secreted by different cells. While CXCL5 secreted by tumor epithelial cells promotes angiogenesis and immune evasion, secretion by CAFs in the microenvironment promotes EMT in epithelial cells(62). Upregulation of EMT has consistently been a poor prognostic factor, predicting for chemoresistance (63). Oxford Classic-EMT (OxC-EMT), a 52 gene signature based on EMT pathways, has the potential to prospectively identify patients likely to have poor prognosis in OxC-EMT high risk (score>0) compared to low risk (score=0) (64,65).

**Fig 4:**
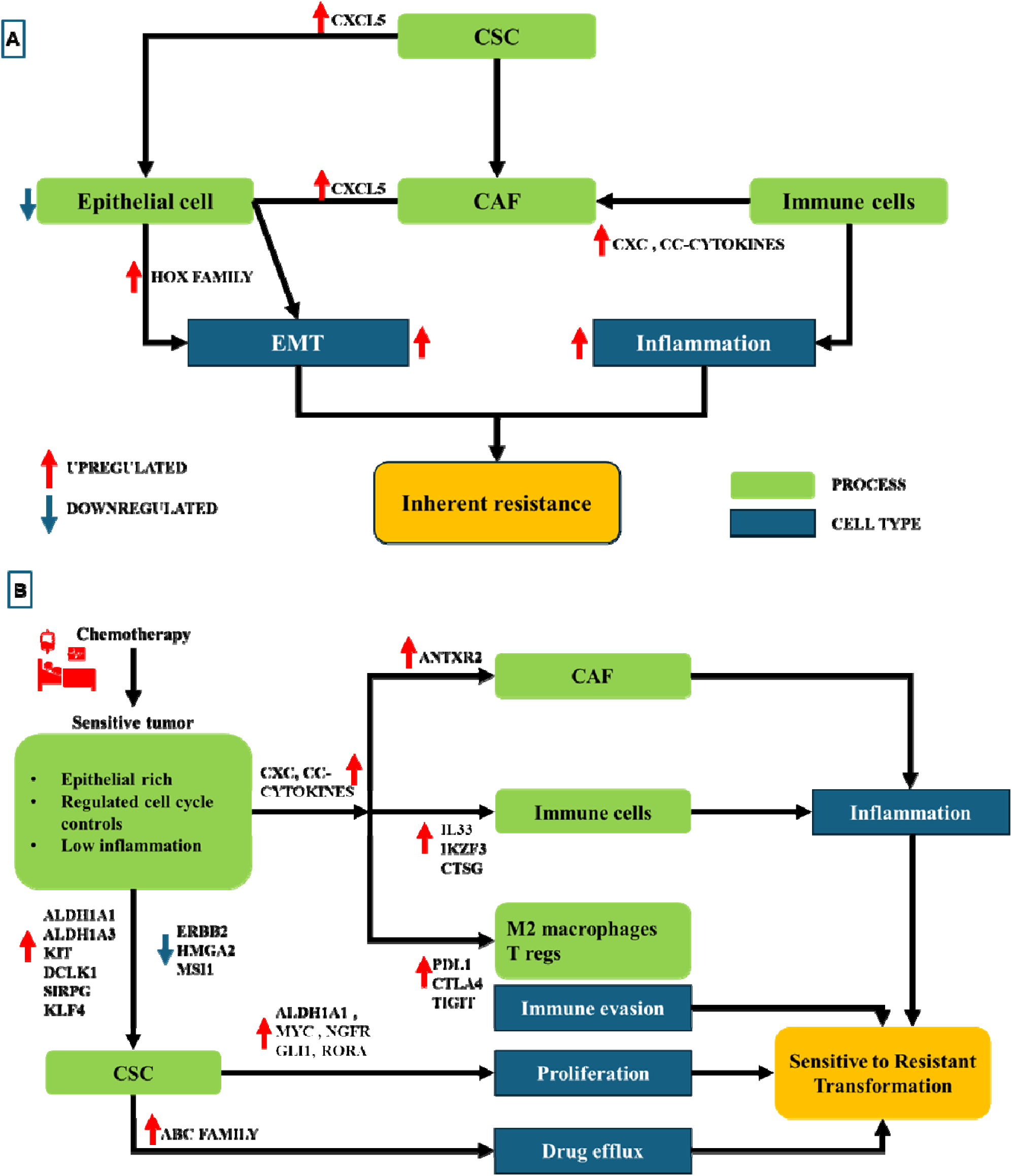
A. Proposed model of inherent resistance. Cancer stem cells influenced by unique regulatory milieu give rise to tumors characterized by lower epithelial to stromal ratio, which is enriched in inflammatory microenvironment. There is high EMT which in combination with inflammatory milieu promotes chemoresistance. B. Proposed model of chemosensitive to resistant transformation after neoadjuvant chemotherapy exposure. Chemotherapy administration on epithelial rich, low inflammatory microenvironment chemosensitive samples leads to CSC proliferation, inflammation, immune evasion and drug efflux leading eventually to transformation of sensitive patients to chemoresistant phenotypes.

Our study suggests that an inherently **chemosensitive** tumor is epithelial-rich, has highly regulated cell cycle controls, and has low inflammation in the microenvironment. On administration of chemotherapy to this phenotype, there is a significant transformation of the tumor microenvironment. CXC and CC-cytokines, and CAFs increase recruitment of monocytes and cytotoxic T cells, causing inflammation, while upregulation of PDL1, CTLA4, TIGIT, and LAG3 promotes immune evasion and exhaustion with infiltration of T Regs and M2 macrophages. Kruppel-like factor4 (KLF4) is a well-known embryonic and cancer stem cell marker, influencing diverse functions in the tumor microenvironment. KLF4 is critical in the development and maturation of multiple immune cells, especially neutrophils, T and B lymphocytes, and monocytes, and plays an important role in normal immunity (66). Significantly, KLF4 induces recruitment of myeloid-derived suppressor cells (MDSCs) *via* CXCL5 activation, which promotes tumor growth in breast cancer models (67). Chemotherapy further upregulates CSC markers, which lead to proliferation (ALDH1A1, KIT, NGFR), immune modulation (CTSG, IKZF3, SIRPG, IL33), and drug efflux pump (ABC family) activation. ABCB1 and ABCG2 are previously known to be predictors of chemoresistance (68) in ovarian cancer patients, and their upregulation in chemosensitive patients on exposure to chemotherapy could contribute to the development of resistance in these patients. The combination of inflammation, immune evasion, CSC reactivation, proliferation, and drug efflux pumps eventually promotes conversion of sensitive-to-resistant phenotype on chemotherapy exposure (**Fig 4B**).

The recurring finding in our analysis is the presence of an immunomodulatory microenvironment resembling the chronic inflammatory phenotype, and lower tumor purity in resistant samples. This is unlike the TCGA transcriptomic subtype analysis, which did not find any significant survival impact of immunoreactive vs other subtypes (69). Our analysis included patients undergoing NACT, which is qualitatively different than the primary surgery phenotype of the TCGA dataset. Chemotherapy-induced inflammation promotes immunogenic cell death, one of the important mechanisms by which chemotherapy exerts cytotoxicity (70). On the other hand, a low CD8+: FOXP3 Tregs ratio predicts immune evasion and poor prognosis in breast cancer treated with NACT (71). In our study, immune microenvironment showed an upregulation of CD8+ and NK cell-mediated immunity counterbalanced by enrichment of Tregs, M2 macrophages, and CAFs, suggesting an immune suppressive microenvironment, a finding previously implicated in ovarian cancer progression (72). The immunosuppressive microenvironment in patients has been shown previously to correlate with poor response to NACT (37). The Wayne group found enhanced T cell-mediated immune response to correlate with chemosensitive disease, though there was no significant change observed in Treg populations, unlike our study (40).

Cytokines promote cancer progression and chemoresistance by various mechanisms, including promoting EMT and inducing a chronic inflammatory milieu (73). On the other hand, cytokines and chemokines also contribute to chemotherapy-induced immunogenic cell death (74). In our study, there was a consistent upregulation of CCLs and CXC cytokines in chemoresistant samples, suggesting their role as a modulator of a pro-tumorigenic microenvironment. CCL2/CCL5 axis increases immune-mediated clearance of circulating liposomal doxorubicin, thereby contributing to ovarian cancer chemoresistance (75). Peritoneal mesothelial cells secrete CCL2, which contributes to the creation of a premetastatic niche promoting ovarian cancer metastasis(76). Mice deficient in mesenchymal stromal cells had reduced involvement of the peritoneal surface by ovarian carcinoma due to reduced CCL2/CCL5 secretion (77). Apart from stroma, CD133+ ovarian CSCs promote invasion and metastatic potential through autocrine CCL5 secretion via NFkB pathway (78) and stimulating EMT (79). In addition, CCL5 promotes the conversion of ovarian CSCs into endothelial cells, leading to angiogenesis (80). High serum CCL2 levels were associated with poor prognosis in nasopharyngeal carcinoma(81) and advanced-stage mesothelioma (82). Circulating CXCL and CCL cytokines could be potential biomarkers for non-invasive identification of chemoresistance in HGSOC owing to their high concentration in human plasma secretome (83).

Despite several studies implying HGSOC to be an immune “hot” tumor, immune checkpoint inhibitors (ICI) have been largely unsuccessful in improving survival (84). CAFs play a major role in modulating immune response and have been shown to reduce survival. Immune CAFs are characterized by low αSMA and high CD74 expression (85), which was the profile seen in Chemosensitive postCT samples in our study. Elevation of CAF population could possibly explain the lack of ICI therapy response in HGSOC. Targeting CAFs has previously been shown to improve T cell-mediated immune response and reduce chemoresistance (86). NACT promotes fibrosis, and ovarian CAFs create a safe niche for CSC-mediated tumor repopulation via non-canonical Wnt pathway (87). Ovarian cancer cells and CAFs show a symbiotic relationship in the tumor microenvironment, with cancer cells influencing conversion of fibroblasts into CAFs, and CAFs on other hand promoting tumor growth and survival while promoting chemoresistance in ovarian cancer through paracrine signaling (88). Among the mesenchymal transcriptomic subtype of HGSOC, CAFs play an immunosuppressive role by promoting T Regs mediated immune evasion (89). In our study, inherent resistant patients had TCGA mesenchymal phenotype with enrichment of CAFs suggesting that immune evasion could be playing a key role in chemoresistance.

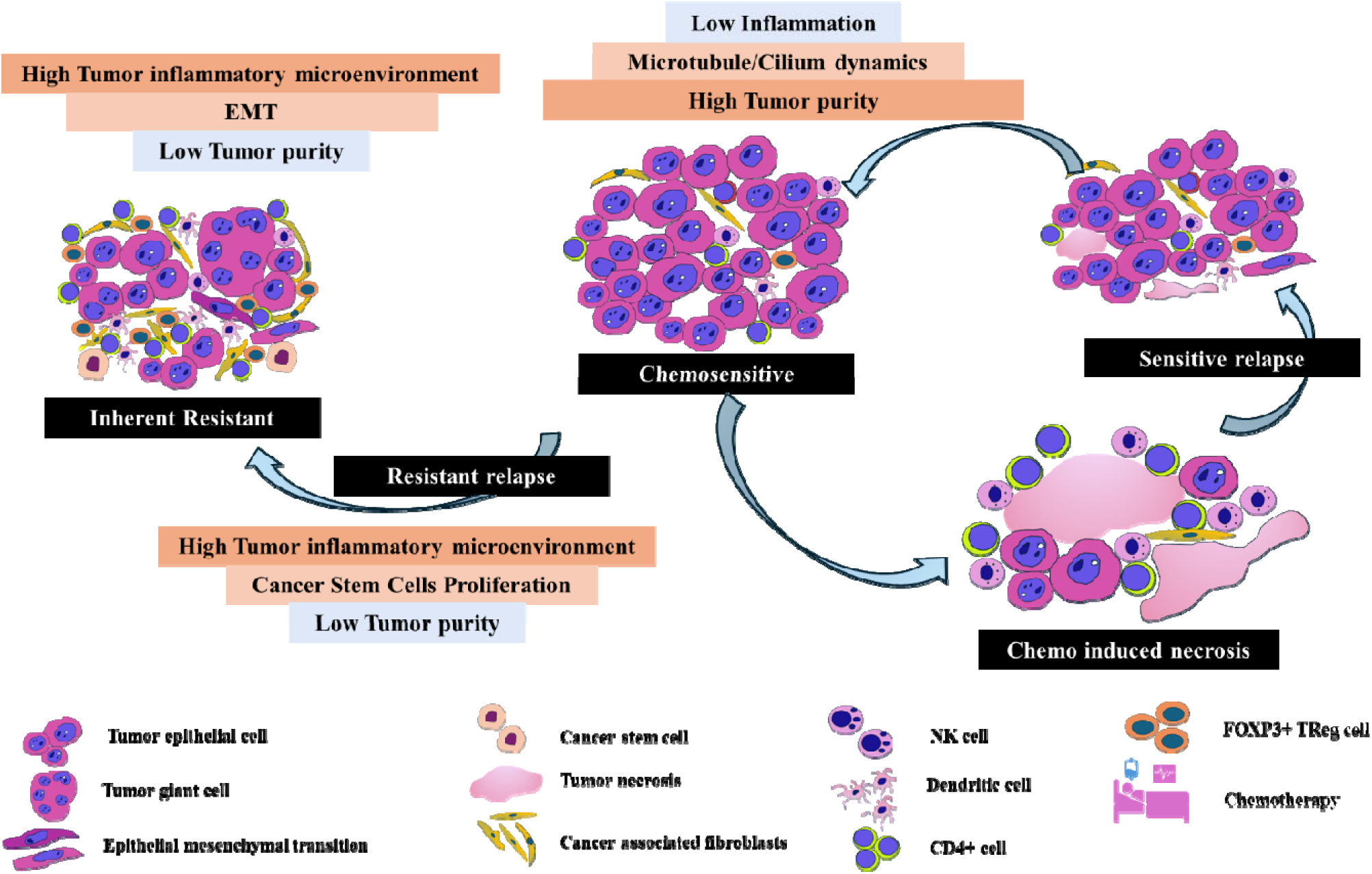

**Graphical summary:** Evolution of chemoresistance in patients on NACT. At the time of presentation, patients are either inherent resistant or sensitive. Inherent (preCT) resistant patients are characterized by high tumor inflammatory microenvironment, epithelial mesenchymal transition (EMT) and low tumor purity. The sensitive patients demonstrate an enriched epithelial phenotype, low inflammation and microtubule / cilium dynamics. On administration of chemotherapy, a subset of sensitive patients show high tumor inflammatory microenvironment, CSC proliferation and reduction in tumor purity, thereby developing acquired resistant phenotype. On the other hand, other subsets undergo tissue necrosis and eventually have platinum sensitive relapse.

### 4.2 Clinical perspective

Neoadjuvant chemotherapy is an acceptable alternative to primary debulking and adjuvant chemotherapy in HGSOC. Our analysis shows that chemotherapy may be provoking an aggressive tumor biology in a subset of patients by promoting CSC activation, inflammation, immune evasion, proliferation, and drug-efflux mechanisms, which might eventually lead to the development of chemoresistance. Identifying these patients could help design novel treatment protocols for them in the future. Immune checkpoint inhibitor (ICI) therapy has not been practice-changing in HGSOC, one of the few solid tumors where it has failed to have a meaningful impact. Perhaps a more nuanced understanding of the immune and stromal microenvironment changes will select the right patients and clinical setting for integration of ICI therapy in clinical regimens.

Our study has several shortcomings. While all the samples were derived from patients and raw data was used to perform the analysis from a relatively homogenous clinical cohort, the findings have not been validated in vitro. We excluded relapsed samples due to lack of paired relapse samples among the different studies included in meta-analysis. Nevertheless, analysis of relapsed samples could help further validate the results obtained. The role of micro-RNAs, other non-coding RNAs, and microbiome was not performed in the current study due to various limitations of data. Changes observed in gene expression may not necessarily be translated into protein expression or function, and hence our study can be considered a hypothesis-generating study. Further *in-vitro,* and clinical validation of questions raised in this study is warranted.

In summary, the role of inflammatory tumor microenvironment appears to be crucial in promoting chemoresistance in advanced HGSOC patient populations. Inflammatory CCL cytokines could potentially be used as serum biomarkers for the prospective identification of chemoresistant patients, thus paving the way for personalized therapy. The administration of NACT promotes sensitive-to-resistant transformation that leads to relapse-chemoresistance cycle. Targeting CAFs could potentially help reduce chemoresistance as well as possibly help sensitize patients to immune-checkpoint inhibitor therapy, especially in chemosensitive patients on NACT protocols. Further studies could help refine our understanding of these processes to manipulate the tumor microenvironment for optimizing survival outcomes.

## Data Availability

All data produced in the present study are available upon reasonable request to the authors.

## 5. ACKNOWLEDGEMENTS

We appreciate Dr. Trivadi S Ganesan (Sri Ramachandra Institute of Higher Education and Research, Chennai, India) for reading this manuscript and providing constructive suggestions. We thank Dr. Mara Artibani (Nuffield Department Women’s & Reproductive Health, University of Oxford, UK) for providing clinical metadata of patients included in GSE162714 dataset. We also thank Dr. Ann-Christin Ostwaldt (Academic coordinator, Sampsa Hautaniemi Lab, University of Helsinki, Finland) for facilitating transfer of data related to GSE173420 dataset through European Genome-phenome Archive (EGA).

