## Supplemental Table for "Does neoadjuvant chemotherapy bias the transcriptomic profile of platinum-sensitive advanced high grade serous ovarian cancer patients towards a resistant phenotype? Findings from a gene expression meta-analysis"

**Supplementary Table : Details of software and versions used in the analysis.**

| **Software** | **Package** | **Version** |
| --- | --- | --- |
| R software |  | 4.3.2 |
| R | tidyverse | 2.0.0 |
| R | ggplot2 | 3.5.2 |
| R | sva | 3.50.0 |
| R | DESeq2 | 1.42.1 |
| R | ggpubr | 0.6.0 |
| R | ggrepel | 0.9.6 |
| R | AnnotationDbi | 1.64.1 |
| R | org.Hs.eg.db | 3.18.0 |
| R | biomaRt | 2.58.2 |
| R | MetaVolcanoR | 1.16.0 |
| R | clusterProfiler | 4.10.1 |
| R | bluster | 1.12.0 |
| R | igraph | 2.1.4 |
| R | instaPrism | 0.1.6 |
| R | pheatmap | 1.0.13 |
| R | immunedeconv | 2.1.0 |
| STRING database | Online tool | https://string-db.org/ |
| Cytoscape | Software | 3.10.3 |
| Cytohubba | Cytoscape plugin | 0.1 |
| GSEA software | Software | 4.3.3 |
